# Mental health challenges and the associated factors in HIV-positive women who have children living with HIV in Indonesia: A qualitative study

**DOI:** 10.1101/2022.05.09.22274877

**Authors:** Nelsensius Klau Fauk, Silvia Merry Maria, Lillian Mwanri, Karen Hawke, Paul Russell Ward

**Affiliations:** Research Centre for Public Health Policy, Torrens University Australia, Adelaide, South Australia, Australia; Institute of Resource Governance and Social Change, Kupang, Indonesia; Medicine Faculty, Duta Wacana Christian University, Yogyakarta, Indonesia; Infectious Disease - Aboriginal Health, South Australian Health and Medical Research Institute

**Keywords:** Mental health challenges, supporting factors, HIV-positive mothers, children living with HIV, Indonesia

## Abstract

Women living with HIV (WLHIV) are vulnerable to various mental health challenges. However, there is a paucity of studies globally and in the Indonesian context that have specifically explored mental health challenges among HIV-positive mothers who also have children living with HIV (CLHIV). This qualitative study explored mental health challenges and the associated factors in HIV-positive mothers with CLHIV in Yogyakarta, Indonesia. In-depth interviews were employed to collect data from the participants (n=23) who were recruited using the snowball sampling technique. A qualitative data analysis framework was used to guide the analysis of the findings. The findings showed that the mothers experienced a range of mental health issues due to their own, and their child’s diagnosis; stress, depression, anxiety, fear, sadness, and guilt. Lack of knowledge about HIV, fear of death, shame, not knowing whom to talk with and what to do after their own HIV diagnosis, and the HIV diagnosis of their children were factors that challenged their mental health. Difficulties in dealing with daily life or social activities of their CLHIV, dilemma in addressing questions and complaints of their CLHIV about HIV treatment, and concerns about the health condition of their CLHIV and how their children cope with any potential negative social impacts also impacted the mothers’ mental health. Social factors such as unsympathetic expressions from friends towards them and their CLHIV and negatively worded religious-related advice from parents and relatives also contributed to their poor mental health. Our findings indicate the need for intervention programs that support HIV-positive mothers and their CLHIV. Future large-scale studies involving HIV-positive mothers with CLHIV in Indonesia and other settings globally are needed to obtain a comprehensive understanding of mental health challenges and the associated factors they face.

## 1. Introduction

The UNAIDS reports an estimated 37.7 million people living with HIV (PLHIV) worldwide in 2020, of which over half (53%)are females aged 15 and over [1]. The same report also shows a high prevalence of HIV infection for females in sub-Saharan Africa, where girls and women living with HIV (WLHIV) account for 63% of total infections in the region [1, 2]. Females in the same age category also represent 30% of the 5.8 million PLHIV across Asia and the Pacific region [1, 2]. In the context of Indonesia, women aged 15 years and over represent 38% of the 427,201 PLHIV in 2021 [3, 4]. Mothers and/or housewives are not only a high risk group for HIV in Indonesia, but also do suffer the most from the impacts of HIV [4]. The Indonesian national AIDS report shows that for the last 10 years, more than one thousand HIV-positive mothers and/or housewives progress to AIDS annually [4]. They are the second highest group of people living with AIDS in the country, comprising 14.3% of the total AIDS cases, following unskilled workers, who account for 16.2% [4].

Existing evidence has suggested that HIV in women in general leads to a range of mental health challenges, including stress, anxiety, depression, sadness and embarrassment [5-10]. The stressors for these mental health issues in WLHIV vary and include advanced stage of HIV infection, weakened physical condition, and the fear of a breach of the confidentiality of their HIV status, which may cause further negative impacts on themselves and their family [9-14]. Other stressors include fear of transmitting HIV to their unborn babies, the burden of being caretaker for their children, concerns about their future and the future of their children, and a lack of resources needed to support their children and family [6, 14-19]. Similarly, the lack of social support, the experience of social rejection and social isolation, avoidance by family members, internalised or perceived stigma [6, 10, 14, 16, 20, 21] and poor economic conditions [8, 11, 17] are also reported determinants for depression, anxiety, fear and worry among WLHIV. Additional factors such as low level of education attainment, cessation of a relationship with a partner or divorce, a partner’s death, family misfortune, family food insecurity and treatment failure [8, 11, 13, 20, 21], are all predictors of negative impacts to mental health challenges.

Despite this, there is a lack of research globally and within Indonesia about HIV-related mental health challenges and the associated factors in HIV-positive mothers who have CLHIV. This study aims to fill this gap in knowledge by interviewing HIV-positive mothers with CLHIV in Yogyakarta, Indonesia. Yogyakarta was chosen as the study setting due to feasibility, familiarity and the potential of undertaking the current study successfully. Exploring these factors is imperative to form an understanding of the lived experience of HIV-positive mothers and their children, to better inform the development of policies and intervention programs that address the complex needs of both the mothers and the CLHIV.

## 2. Methods and analysis

A qualitative methodology was used to explore mental health impacts of HIV on HIV-positive mothers who have CLHIV. The study focused on understanding mental health challenges and the associated factors they faced following their own and their children’s HIV diagnosis. We recruited the participants using the snowball sampling technique. Initially, we solicited the help from the head of an HIV clinic and a companion of PLHIV, who is also HIV-positive, in the study setting. Both agreed to help distribute the study information sheets containing contact details of the researchers to potential participants. The information sheets were then posted on the information board at the clinic by the clinic receptionist and on the WhatsApp group of PLHIV by the companion of PLHIV. Potential participants who contacted the researchers and confirmed their participation were recruited for an interview. After the interviews, initial participants were also asked for help to distribute the study information sheets to their eligible friends who might be willing to participate in this study. None of them who texted or called to confirm their participation withdrew from the study. They were recruited based on several inclusion criteria: one had to be 18 years old or above, and a mother living with HIV who has a child or children living with HIV. Finally, we recruited and interviewed 23 participants. The participants’ age ranged from 24 to 43 years old. The majority graduated from senior and junior high school and were engaged in different kinds of professions, while five were housewives and three were unemployed. They had all been living with HIV for different periods of time. All participants and their CLHIV were on antiretroviral therapy (ART) when the interviews were conducted. Each participant had only one child living with HIV.

In-depth interviews were carried out to collect the data from the participants in Yogyakarta, Indonesia. Interviews were conducted face-to-face and online using WhatsApp video calls. These modes of interview and interview times were decided based on the preference of the participants. Interviews explored participants’ views and experiences of mental health challenges they faced following their HIV diagnosis; their views and experiences of how their children’s HIV diagnosis influenced their mental health condition; their experience of dealing with or managing social activities, social life, health condition and treatment of their CLHIV and how these factors influenced their mental health condition; the interviews also explored their views about the influence of their social relations with families, relatives and friends on their mental health condition. The duration of the interviews was approximately 40 to 60 minutes, with only an interviewer and each participant present during the interviews. Interviews were conducted in Bahasa Indonesia, the primary language of the interviewers (NKF, MSM) and the participants, and were recorded. Sometimes notes were also taken during the interviews, if felt necessary. Interviews ceased when we felt that data had been rich enough to address the research questions and aim, and that data saturation had been reached and no new information was obtained from the responses of the last few participants. At end of the interviews, we offered each participant an opportunity to read and correct the information they had provided after the transcription of the recordings, but none took the opportunity. The participants were assured that information they provided would remain confidential and anonymous, and a pseudonym was assigned to each interview transcript. They were also advised prior to the interviews that they had the rights to withdraw from the study for any reason, at any time. Each participant signed an informed consent to indicate their willingness to participate and returned it in person or via WhatsApp prior to or on the interview day.

Interview recordings were transcribed verbatim for analysis. Transcription occurred alongside the data collection process which enabled direct integration of notes taken during the interviews into each transcript. Informed by Ritchie and Spencer’s qualitative data analysis framework, five steps were undertaken throughout data analysis process: 1) familiarisation with the data, 2) identification of a thematic framework, 3) indexing the data, 4) charting the data, and 5) mapping and interpretation of the data [22, 23]. Familiarisation with the data was performed by reading each individual transcript repeatedly. While reading the transcripts, the participants’ narratives were broken down into chunks of information, which were then labelled and commented. Key issues, concepts and ideas that emerged from the transcripts were then identified and listed, and then used to form a thematic framework. Data extracts in each individual transcript were indexed or coded entirely (open coding). This was followed by the identification of similar or redundant codes which were then collated (close coding). Codes that formed a logical pattern to explain a theme were grouped together under the theme or sub-theme. All themes and their codes which had been created in the previous steps were then reorganised in a summary of chart for the purpose of comparison across the interviews. Finally, the entire data were mapped and interpreted. This data analysis was non-linear and an iterative process which involved changing and refining codes and themes before we agreed upon the final themes presented in this paper. The steps undertaken in this analysis helped us manage the data in a structured and coherent manner and enhanced the rigour, transparency and validity of the analytical process [22, 23].

Ethics approval for this study was obtained from Health Research Ethics Committee, Duta Wacana Christian University, Indonesia.

## 3. Results

### 3.1 The experience of mental health challenges

Mental health challenges were experienced by all the study participants after their HIV diagnosis. They described that it led to *“extreme stress and depression for a few years”* (Ayu) and *“feeling sad, afraid and anxious every day at the beginning of the diagnosis”* (Indah). Lack of knowledge about HIV, fear of death, shame, and not knowing anybody to talk with and what to do following the diagnosis were some of the contributing factors for the feelings of stress, depression, anxiety, fear, sadness they experienced. The following narrative of a participant who had been diagnosed with HIV for nearly five years illustrates how such factors influenced her feelings after her HIV diagnosis:

> *“After being diagnosed with HIV what I could do was crying. I was very stressed, sad, scared, all mixed up. I didn’t know anything at all about HIV, haven’t heard of it before. I was very scared because I thought I was going to die soon. I didn’t know whom to ask for help, whom to talk with about this problem. I didn’t know what to do. I didn’t dare to seek help from other people because I was ashamed if people know my HIV status. I remember I was so depressed. So, for at least two years I struggled with those feelings myself” (Julia)*.

The HIV diagnosis of their child was another factor that impacted their mental health significantly. They acknowledged that it had increased the burden on them and put them into a situation where they often felt guilty *“Since my second child tested positive with HIV, I feel very much guilty. I often look at my child and cry, and say ‘sorry mom has made you suffer from this [HIV]* (Anti)*”*. They also experienced prolonged worry *“I am worried all the time about my child since my child tested positive last year. I have been thinking too much”* (Dewi). The stories of the participants also indicated that once their child was diagnosed, there was a definite shift in their focus, with much of their attention and thoughts going to their CLHIV. The following narrative of a participant who has a child living with HIV depicts such experiences:

> *“After I received the [HIV] test result of my child, I told myself that I must be strong and focus on the life of my child. But you know what, I feel the burden is even heavier. These feelings: afraid, sad, worried, stressed, guilty, don’t go away. I feel these up to now, but not because I am caring about myself. It is all about my child. I think a lot about my child and put all my attention and focus on my child. I fear any negative impacts my child may face in the future due to this [HIV] and it makes me sad and stressed out. It is my mistake that I transmitted HIV to my child, my child doesn’t deserve this. The more I think about the situation and future of my child the more worried and stressed I get. I have been trying to live a normal life without thinking too much but these thoughts and feelings never go away”* (Jeane).

### 3.2 Daily life challenges associated with their children’s condition of living with HIV

Dealing with daily life or social activities of their CLHIV was a significant challenge for the mothers. They described that their children’s daily activities, such as going to school, mingling with friends, and engaging extracurricular activities with other students including school excursions, picnics, and sport, increased their fear and worry about the possibility of their children’s HIV status being disclosed or discovered. Yeni, who had a child living with HIV, stated *“I am worried every day because my child often mingles and plays football with other kids. On the one hand, I do not feel right to forbid my child to play with other kids, but on the other hand I am worried if other kids find out about HIV status of my child”*. Similarly, Enda who also has a child living with HIV described *“I do not feel calm and am always worried every day I walk my child to school or when my child and other kids work on their homework at other kids’ house. I am afraid if other kids or their parents are suspicious and find out that my child has HIV”*. Such over thinking and fear experienced by the mothers seemed to be influenced by their projection of negative impacts that could happen to their children disclosure to their friends and/or other people of their HIV status. The following quote illustrates such assertions:

> *“My child is now in junior high school. Sometimes they [her child and other kids] go out together. They do their homework together. Sometimes, my child goes to other kids’ place, or other kids come to our house. I am happy that my child has friends but deep inside I have always been worried and afraid if my child accidently tells other kids about the condition [her child’s HIV status]. I am scared thinking of any possible negative impacts that could happen to my child if other people know about this [her child’s HIV status]. Other kids may avoid or reject my child and this could make my child down, stressed and unwilling to go to school. All the activities my child engages in always bring me these kinds of thoughts and fears. Sometimes, I want to limit my child’s activities but I can’t do that because I see my child is happy doing those activities. It hasn’t been easy for me to deal with my child’s daily life and activities…*.*”* (Betty).

Most mothers in the study acknowledged that their CLHIV were not aware or had not been informed about their HIV status because of their young age. This complicated the mothers’ experience of living with HIV and they found it very challenging managing their children’s health condition, especially when their children disliked medications and questioned why they needed to take them regularly. For example, the children would ask: *“Why should I take this [ART] medicine every day?”* (Silvia), *“Can I skip taking the medicine today?”* (Astri), *“I don’t want to take the medicine”* (Retno). This questioning, complaining and rejection by their children complicated the prevailing situations, compounding the impact of HIV on mental health of these women:

> *“I just don’t know how to tell my child [about his HIV status] because my child is still little, eight years old. My child can’t understand the complex issue around HIV and may tell other kids about it [her child’s HIV status] without knowing the negative consequences that may happen. No one can guarantee that my child will keep it secret. It has been very stressful for me because my child keeps on asking why taking the medicine every day. My child complains a lot and sometimes cries and rejects to take the medicines. I feel guilty for what my child is going through and will be facing in the future”* (Fatima).

Concerns about what action they need to take if their children get sick and how to prepare their children to deal with their health condition and the surrounding social environment in the future were also factors that made them feel stressed and scared. Yanri, who has a child living with HIV, described *“We [the woman and her spouse] have always been trying to prepare ourselves and think of what to do if our child falls ill, and this is stressful considering my situation of not having a permanent job or purely relying on my spouse”*. She continued *“I am also concerned with how my child will deal with the condition [HIV-positive status] and interact with other people in the future. I want to prepare my child for these and I feel scared”*. Such concerns seemed to also be influenced by their unstable economic or financial conditions and negative treatments towards their children or other children living with HIV by uninfected people within families, communities and healthcare settings:

> *“I am concerned with my child’s health condition. I see that my child is not very strong and can get sick anytime. What makes me stressed out is that I don’t have a job, no income at all. I always think of what I am going to do if my child gets sick or is admitted to hospital. So far, we [the woman and her child] rely on financial support from my parents. The situation I am in now makes me worried. To be honest I am scared deep inside. I have just submitted an application for a job a few days ago, hopefully I get accepted”* (Kana).
>
> *“We [the woman and her HIV-positive child] experienced negative treatment once at the hospital. We were not served; my child was not allowed to use the toilet at the hospital. It was three years ago, my child was four years old so he didn’t really understand. That is why I am concerned with how to prepare my child to face such kinds of negative treatments. There are many kids [living with HIV] who receive negative treatments from their friends at school or within communities. I don’t want my child to feel broken if these happen, that is why I am worried”* (Ratih).

### 3.3 Social factors: unsympathetic expressions and comments from others

Social factors such as unsympathetic expressions and comments from friends towards participants’ children living with HIV negatively impacted their mental health condition. A few participants, who acknowledged having told their close friends about their own and their children’s HIV status, described that some of their friends seemed to be trying to show caring attitudes or attentions towards CLHIV, but their expressions and comments sometimes made them feel guilty, attacked, and stressed. The quote presented below reflects such kinds of expressions or comments they received from their friends:

> *“There are some close friends of mine whom I have told about my own and my child’s situation because they have been nice to us. But sometimes their words which seem to reflect their sympathy for my child but actually pierce my heart. For example, one of them said to my child ‘You are a strong child, keep your spirit up, take medicine. It’s not your fault, you should be fine’. In other words, it’s your mother’s fault. I feel accused and guilty. I sometimes feel stressed out when I remember other people’s words in this tone. After I heard what they said I try to keep distance from them, we are not that close anymore”* (Shinta).

Similarly, unsympathetic comments from friends towards the participants themselves were acknowledged as having an influence of their mental health condition. The following story of a woman, who acknowledged contracting HIV from her ex-boyfriend prior to her marriage with her current spouse, reflects unsympathetic comments of her friends which made her feel shocked and depressed:

> *“Some of my friends, who know that we [the woman and her child] are HIV-positive, have negative comments about me. Some said ‘This is a consequence of her own actions. She had sex with several men before marriage, that is why she got HIV. It’s a pity that her little child must also bear the consequences’. When a very close friend of mine told me this [what her other friends said about her], I was shocked and depressed because I never thought that they had such a negative view about me”* (Ima).

### 3.4 Religious advice from family members

Religious advice received from parents and relatives were also reported by some participants to bring back their memories of some difficult past experiences they had gone through. Some described that their parents’ religious advice reminded them of the mistakes they had committed in the past, and these were acknowledged as not helping but making them feel guilty, angry and stressed. The following story from a participant reflects such experiences:

> *“My parents always give me advices. I remember they said ‘You have made mistakes and what you are now going through is a warning from God for you. Now you should try to get closer to God…*.*’. I understand what they mean is that I have been guilty of having children before marriage or not having a husband. This is wrong because it is not in accordance with our [Islamic] religious teachings. As a result of my mistakes, my child and I got HIV. This seems like a good advice but it makes me feel guilty and stressed out. Advice from parents sometimes makes me get mad at myself and what I have done”* (Rini).

Similar comments were raised by another participant who acknowledged to engage in injecting drug use and group sex prior to her marriage. She reported frequently receiving religious advice from her relatives, but felt accused, disappointed, and stressed:

> *“Some of my relatives care about me and my child. They often support us financially and give me advice to stay strong: ‘You have to pray a lot, God listens to people who are guilty but want to repent for their mistakes and avoid sins. This is also for the good of your child because your child doesn’t deserve this [infection]’. Sometimes I take my time and think about the advice they have given me, and I feel like they are accusing me of what I did in the past. That’s why I don’t really like to visit them anymore in their house. It has been a while we don’t meet each other”* (Lina).

## 4. Discussion

Women diagnosed with HIV infection are vulnerable to a range of negative impacts, including mental health issues. This study highlights that HIV-positive mothers experience various mental health challenges, including stress, depression, anxiety, fear, sadness and feelings of guilt, that are line with findings of previous studies [6-10, 24]. As is the experience of many other WLHIV in general [9, 13, 25-28], a fear of death and shame due to HIV infection were two prevalent factors that contributed to mental health challenges facing these mothers.

Previous studies have found that a lack of knowledge about HIV majorly contributes to feelings of stress and fear among WLHIV following their HIV diagnosis [20, 29]. This is also apparent in our findings, mothers reported not understanding HIV and feeling there was a real lack of education and resource for them, which made them feel stressed, depressed, anxious and fearful, particularly during the early stage of their infection and when they found out that their child was infected. What our study adds to current literature is that mothers talked about not knowing whom to confide in and who to go to for information. This led to feelings of isolation, and seemed to prevent them from taking necessary actions in such a challenging circumstance. In addition, there is strong indication from our findings that these mothers were not informed or aware of any available social and health supports they could access to help themselves cope with mental health challenges and other detrimental impacts they experienced following their HIV diagnosis. This reflects how important it is to have freely available HIV-related health literacy for WLHIV and mothers of CLHIV, so they can make critical health decision [30].

Our study also reports novel findings on factors associated with mental health challenges facing HIV-positive mothers, which have not been explored previously [6-10, 24]. For these mothers, having a child diagnosed with HIV doubled the existing burden as it shifted their focus and attention from themselves to their child’s health, and made them feel even more stressed, scared and uncertain. This enriches current evidence about double burdens PLHIV experience, from things including; co-infection, malnutrition and food insecurity [31, 32]. Mothering CLHIV requires an enormous amount of love, devotion, and care, and takes an emotional and physical toll, especially under difficult circumstances when support is scarce. On top of this, mothers constantly worry and live with anxiety, with intrusive thoughts about themselves or their children dying, blaming themselves and feeling chronic guilt for infecting their child unintentionally [33]. Our study also identified that mothers were worried about what the future holds for their child, and experienced difficulties in dealing with daily life or social activities of their CLHIV. The mothers often projected the possibility of negative consequences their CLHIV may encounter if their HIV status was made known to others. Such concerns are valid; children’s involvement in various social activities with their friends increase the possibility of their HIV status being accidently disclosed to or discovered by their friends, which could lead to negative social consequences, such as stigma and discrimination. Such concerns are underpinned by previous experiences of negative treatments towards their children and other PLHIV by non-infected people with families, communities and healthcare settings, which have also been reported in previous studies in different settings globally [10, 16, 20, 21, 25, 26, 34].

The mothers in our study also reported the difficulties and dilemma they faced when it came to dealing with questions and complaints from their CLHIV about their health condition and routine ART. Similarly, concerns about the possibility of future healthcare expenses for their CLHIV’s health needs due to poor economic and financial situations, were also supporting factors for feelings of stress and fear among these mothers. These are consistent with the previous findings [8, 11, 17, 35] which have reported poor economic condition and healthcare expenses as determinants of depression, anxiety, fear and worry among WLHIV. It is therefore apparent that economic or financial factors played an important role in influencing these mothers’ response to their children’s health condition, which in turn exacerbated their mental health state.

Findings from previous studies have reported the important influence of social factors such as stigma and discrimination against WLHIV, reflected in social rejection, social isolation, avoidance by others, on their mental health condition [6, 10, 14, 16, 20, 21]. Our study adds further evidence on social factors such as unsympathetic expressions and comments from others and religious-related advice from parents and relatives which contained negative words as contributing factors for various mental health issues experienced by the participants. It is also plausible to argue that a lack of empathy from others, and negative words the mothers receive may also lead to their withdrawal from social relationships with friends and relatives, and to them missing social support from their close ones. Negative attitudes of and lack of support from families and friends have been reported as having significant influence on access to social support and health care services by PLHIV [25, 26, 34, 36-38]. These findings demonstrate vulnerability in these participants and the role that complex socio-cultural factors play in impacting the mental health and wellbeing of women and their children living with HIV. These findings are critical and they do inform the need to address them in order to improve health and wellbeing of women and children, including families and societies that live with HIV.

### 4.1 Study limitations and strengths

There are some limitations of the current study that need to be considered in interpreting its findings. The use of snowball sampling technique for the recruitment of the participants and the recruitment through an HIV clinic and WhatsApp group of PLHIV may have led to the recruitment of participants from the same networks of mothers who were accessing HIV care services for them and their children. These may have led to the under-sampling of mothers who were not in the healthcare service networks who may have different stories to tell. Thus, the findings presented in this study mainly reflect experiences of a specific group of mothers. However, the strength of this study is that, to best of our knowledge, it is an initial study that specifically involved HIV-positive mothers who have CLHIV in the context of Indonesia. The current findings have significant implications for the health sector in Yogyakarta and Indonesia as a whole. Mothers and/or housewives living with HIV represent a significant percentage of PLHIV in Indonesia, thus development of policies and interventions that address the needs of both this specific group of mothers and their children who are also living with HIV in the country and other similar settings globally is necessary and highly recommended.

## 5. Conclusions

This study reports a range of mental health challenges, such as stress, depression, anxiety, fear, sadness and the feelings of guilty experienced by HIV-positive mothers who have CLHIV in Yogyakarta, Indonesia. Mothers in Yogyakarta do not feel supported in coming to terms with their HIV diagnosis, or that of their child, nor do they feel they have access to adequate social, financial, and medical support. Fear of stigma and discrimination is a prevalent issue for WLHIV and CLHIV, which has very negative impacts on mental health, especially for the mothers, Currently, there is very limited support for PLHIV in general, and even more so for WLHIV and CLHIV. There is urgent need for further research, and targeted programs that support the needs of HIV-positive mothers who have CLHIV. These may include education or counselling programs for how to deal with HIV-related negative feelings, manage social interactions, and activities of their CLHIV, and address their HIV-related questions and complaints. Financial support through provision of small-scale business capital for example, would also be ideal. Future large-scale studies involving HIV-positive mothers with CLHIV in Indonesia are warranted, if we are to gain a comprehensive understanding of their collective experiences and resulting mental health challenges, so the right supports can be put into place for the women and their children to thrive and feel supported.

## Data Availability

All data produced in the present study are available upon reasonable request to the authors

## Acknowledgments

We would like to thank the participants who had spent their time to voluntarily take part in the interview and provided us with valuable information.

## Author contributions

NKF was involved in conceptualisation, project administration, investigation, and in developing the methodology, conducting formal analysis and writing the original draft of the paper. MSM was involved in project administration and investigation. PRW, KH and LM were involved in conceptualisation, developing the methodology, reviewing and editing the paper critically for important intellectual content.

